# Achieving Inclusive Healthcare through Integrating Education and Research with AI and Personalized Curricula

**DOI:** 10.1101/2024.07.31.24311182

**Authors:** Amir Bahmani, Kexin Cha, Arash Alavi, Amit Dixit, Antony Ross, Ryan Park, Francesca Goncalves, Shirley Ma, Paul Saxman, Ramesh Nair, Ramin Akhavan-Sarraf, Xin Zhou, Meng Wang, Kévin Contrepois, Jennifer Li-Pook-Than, Emma Monte, David Jose Florez Rodriguez, Jaslene Lai, Mohan Babu, Abtin Tondar, Sophia Miryam Schüssler-Fiorenza Rose, Ilya Akbari, Xinyue Zhang, Kritika Yegnashankaran, Joseph Yracheta, Kali Dale, Alison Derbenwick Miller, Scott Edmiston, Eva M McGhee, Camille Nebeker, Joseph C. Wu, Anshul Kundaje, Michael Snyder

## Abstract

**Background:** Precision medicine promises significant health benefits but faces challenges such as complex data management and analytics, interdisciplinary collaboration, and education of researchers, healthcare professionals, and participants. Addressing these needs requires the integration of computational experts, engineers, designers, and healthcare professionals to develop user-friendly systems and shared terminologies. The widespread adoption of large language models (LLMs) such as Generative Pretrained Transformer (GPT) and Claude highlights the importance of making complex data accessible to non-specialists.

**Methods:** We evaluated the Stanford Data Ocean (SDO) precision medicine training program’s learning outcomes, AI Tutor performance, and learner satisfaction by assessing self-rated competency on key learning objectives through pre- and post-learning surveys, along with formative and summative assessment completion rates. We also analyzed AI Tutor accuracy and learners’ self-reported satisfaction, and post-program academic and career impacts. Additionally, we demonstrated the capabilities of the AI Data Visualization tool.

**Results:** SDO demonstrates the ability to improve learning outcomes for learners from broad educational and socioeconomic backgrounds with the support of the AI Tutor. The AI Data Visualization tool enables learners to interpret multi-omics and wearable data and replicate research findings.

**Conclusions:** SDO strives to mitigate challenges in precision medicine through a scalable, cloud-based platform that supports data management for various data types, advanced research, and personalized learning. SDO provides AI tutors and AI-powered data visualization tools to enhance educational and research outcomes and make data analysis accessible to users from broad educational backgrounds. By extending engagement and cutting-edge research capabilities globally, SDO particularly benefits economically disadvantaged and historically marginalized communities, fostering interdisciplinary biomedical research and bridging the gap between education and practical application in the biomedical field.

**Plain Language Summary:** Precision medicine is the use of various types of health data specific to an individual to improve disease prevention, diagnosis, or treatment. We used artificial intelligence to build a precision medicine learning platform for clinicians and researchers in training. Students in 95 countries accessed the platform and found it helpful. It could be particularly helpful for training students in low- and middle-income countries.

## Introduction

Precision medicine utilizes comprehensive health data to facilitate individualized disease prevention, diagnosis, and treatment by accounting for distinct biological, lifestyle, and environmental differences^1^. The process of extracting valuable insights from health data necessitates bioinformatics expertise, access to low-latency systems for secure and scalable data collection, and efficient processing and storage of vast volumes of multi-modal data. However, the high costs of acquiring such expertise and computing resources confine precision medicine advancements to well-funded institutions in high-income countries (HICs), thereby perpetuating disparities in biomedical research, disease diagnosis, and treatment in low- and middle-income countries (LMICs)^2,3^.

Training precision medicine professionals in a wide range of communities not only fosters inter-regional research collaborations that pool broad expertise and financial resources but crucially enhances local healthcare outcomes, especially in under-resourced communities. By empowering communities with skilled professionals, more tailored and effective health interventions will be enabled, directly addressing their unique health challenges. This approach not only generates significant amounts of data and leads to high-impact publications^4^, but more importantly, it translates into tangible improvements in personal health, ensuring that the primary goal of such training is to enhance the well-being of community members.

By incorporating diverse data and facilitating extensive knowledge exchange, noteworthy collaborations have proven to accelerate biomedical discoveries, such as the Encyclopedia of DNA Elements (ENCODE)^5^, the Human Microbiome Project^6^, the International Cancer Genome Consortium (ICGC)^7^, the Human Heredity and Health in Africa (H3Africa) Initiative^8^, and the Global Alliance for Genomics and Health (GA4GH)^9^. By equipping professionals in underrepresented communities with precision medicine training, we hope to better serve them as well as directly address local data deficits and ethical challenges, thereby enhancing health outcomes. Moreover, the study of underrepresented groups is expected to generate new knowledge. For example, the H3Africa Initiative revealed more than 3 million new genetic variants relevant to viral immunity, DNA repair, and metabolism from data of 426 people across 50 African ethnolinguistic groups^10^. Since only 2% of data in genome-wide association studies (GWAS) was from African populations, few research discoveries are specific to these populations^11^. The inclusion of under-represented groups in scientific research was conducted in compliance with U.S. nondiscrimination laws, including Title VI of the Civil Rights Act of 1964. In addition, research that involves American Indian/Alaska Native (AI/AN) peoples must respect tribal self-determination and data-governance principles affirmed in the Indian Self-Determination and Education Assistance Act of 1975. For AI/AN communities, the data deficit is partially due to mistrust stemming from historical and recent research practices^12,13^. However, prioritizing the communities’ needs and incorporating tribal governance in the research process have led to recent successful collaborations^14^.

In addition, the growing bioinformatics and engineering demand in precision medicine calls for effective training of non-life science professionals to contribute to large-scale initiatives, such as the Molecular Transducers of Physical Activity Consortium (MoTrPAC)^15^, Human BioMolecular Atlas Program (HuBMAP)^16^, Human Tumor Atlas Network (HTAN)^17^, Bridge to Artificial Intelligence (Bridge2AI)^18^, Human Microbiome Project (HMP)^19^, and Genotype-Tissue Expression (GTEx) project^20^. Large language models (LLMs) have the potential to become the propeller to deliver personalized education at scale, and to enable patients to gain actionable insights from healthcare data^21–24^. Studies have shown that personalized learning can significantly increase student engagement and satisfaction, leading to improved learning outcomes^25–29^.

In order to enable all communities to become active contributors to precision medicine research and improve community health, we designed and implemented the Stanford Data Ocean (SDO) as a cloud-based, serverless, LLMs-powered platform for researchers and learners to seamlessly access and analyze large biomedical datasets, including public datasets—such as multi-omics and wearables. SDO also offers precision medicine certificate training. Since June 2023, SDO has offered 3,594 scholarships to low and middle income learners in all 50 states in the U.S. and 94 countries so that they have the opportunity to receive the Stanford Genetics certifications in Bioinformatics and AI/ML at no cost. The platform provides free education for learners whose annual income is under $70,000 USD/year, referencing the 2022 United States median income, $74,580 USD/year. In addition, the platform has been collaborating with biomedical education and research leaders in the underrepresented communities, such as Martin Luther King Jr. Community Healthcare Hospital and Native BioData Consortium, to invite students and mentees in the communities to join the programs.

Through the comprehensive curriculum and LLM-enabled AI Tutor and AI Data Visualization tool, SDO supports learners from broad academic and socio-economic backgrounds to develop essential skills in computing and bioinformatics, equipping them for advanced careers in data science. Notably, 22.8% of certified students report the program positively impacts them securing positions within STEM fields (Supplementary Data 1).

## Methods

### Scalable, Secure, and Sustainable Platform

SDO leverages containerization and virtual machines (VMs) to enhance the learning experience. Containers create a stateless environment that facilitates quick setup and disposal, whereas VMs support a stateful environment necessary for continuous operations and complex computations. Together, these technologies ensure uninterrupted access to educational and research content, even amid system changes (see Supplementary Figure 1a). Additionally, the platform’s microservice architecture enhances scalability and security while reducing management overhead. A front-end cluster manages user access and coordinates the operation of VMs and containers, while a back-end cluster safeguards sensitive data and hosts applications. Furthermore, real-time monitoring tools ensure the performance remains optimal and compliant with HIPAA standards for privacy and security^30,31^.

To further strengthen security, SDO is deployed within secure environments such as Amazon Bedrock^32^, Azure OpenAI Service^33^, or GCP Vertex AI^34^ for pre-trained models, with AWS Bedrock serving as the primary environment (see Supplementary Figure 1b. and 1c). This setup gives organizations control over their data and infrastructure in a monitored environment. Using third-party models under stringent data privacy agreements offers an additional layer of protection against data exposure risks. For example, OpenAI approved our request to not use SDO content to train their models. Moreover, sharing data summaries rather than complete datasets minimizes the risk of sensitive information leakage, a technique SDO utilizes in its AI visualization tool.

Security models designed for LLMs incorporate secure deployment environments, data access controls, and test-time defenses^35^ to safeguard data integrity and protection. Collectively, these strategies, along with robust test-time defenses that analyze user prompts and monitor LLM outputs to ensure safety and appropriateness, establish a robust security framework, enabling organizations to confidently utilize LLMs while upholding the highest standards of data protection^35–37^.

Moreover, SDO also standardizes modules such as notebooks and datasets to enhance accessibility and integration for researchers and learners from broad educational backgrounds. This standardization promotes consistency and reproducibility in scientific research while continuously updating resources to keep pace with technological advances, supporting the sustainable development of bioinformatics education. SDO, designed as a comprehensive platform, adheres to FAIR principles—ensuring data is findable, accessible, interoperable, and reusable—thus improving resource efficiency and impact^38^. It also facilitates environmental sustainability in precision medicine through shared data resources. By integrating scientific papers as research modules, such as the NightSignal algorithm^39^, into its ecosystem, SDO lowers entry barriers and simplifies learning for beginners while emphasizing the critical, resource-intensive task of curating and cleaning datasets essential for personalized medicine.

### AI Tutor

Our LLM-powered AI Tutor, developed on the SDO platform, democratizes private tutoring for students who cannot afford or allocate time for traditional methods. This AI Tutor (see Supplementary Figure 1b.) is specialized in questions pertinent to bioinformatics. It operates by receiving student inquiries, applying embedding techniques to identify the most relevant content within SDO, and using prompt engineering to generate pertinent responses.

### LLM-based Data Visualization

As LLMs continue to evolve, automatic data visualization is becoming increasingly prevalent. Systems such as LIDA^40^ and Amazon Q^41^ exemplify a multi-stage generation approach, showcasing how well-orchestrated pipelines that include LLMs effectively address various challenges. However, its generalizability suffers from restricted data intake formats, often limited to common spreadsheet file types and not suitable for multi-omics datasets.

To address these shortcomings, we leveraged some of LIDA’s capabilities such as UI and goal generation and introduced a novel grammar-agnostic data visualization component within the SDO framework (Supplementary Figure. 1c). After uploading a public dataset, the learner will receive a summary describing its contents. They can then choose to create a visualization using an existing template or generate a new one by selecting a research goal suggested by the AI Data Visualization tool. Once the visualization is generated, the learner can copy the underlying code and modify it as needed. If further refinements are required, they can enter a prompt to adjust the visualization accordingly. Additionally, learners have the flexibility to generate visualizations using Python or R, with support for LLM options such as GPT, Gemini, and Claude. Our component transcends the limitations of prior systems by incorporating robust mechanisms for: 1) Multi-modal analysis: It seamlessly integrates insights from diverse datasets, enabling comprehensive data exploration; 2) Enhanced data format support: It ingests a broader range of data formats beyond conventional spreadsheets (e.g.,.xls,.csv) to include geospatial information (maps), compressed archives (zips), and even image data; 3) Automatic error handling: Our system proactively identifies and addresses potential issues during the visualization generation process. This includes situations when model capacity is exceeded due to large context size or when the LLM fails to produce valid executable code. This expanded data intake empowers researchers to conduct richer analyses and unlock hidden patterns across a broader spectrum of information sources, including not just traditional tabular data but also spatial relationships, archived content, and potentially valuable visual information; and 4) Supporting multiple programming languages: Our platform supports Python and R, enabling a broader range of bioinformatics workflows and user preferences. For example, many researchers use R’s Seurat package for single-cell RNA-seq data analysis, while others prefer Python’s Scanpy for similar tasks.

The initial step involves extracting metadata from uploaded datasets, which includes critical details such as column names, data types, and record counts. Following metadata extraction, a representative sample of the dataset is taken to facilitate quick data analysis. Summarization then generates concise descriptions, including statistical summaries and key pattern identifications. Semantic typing categorizes the data into meaningful types, which is essential for selecting appropriate visualization techniques. The selection and instantiation of prompt templates guide the LLM in generating necessary codes or descriptions for creating visualizations. Mechanisms are in place to handle errors, and the process includes steps to iterate through the inference process, maintaining memory and context to complete the output. In scenarios involving multi-modal contexts, the pipeline accommodates the complexity of handling multiple datasets, ensuring visualizations are based on relevant and accurate data. This structured approach to data visualization enhances the effectiveness of data analysis, making complex information more accessible and actionable.

### Statistics and Reproducibility

We compared the skills confidence level rated by 1,495 graduated learners and their satisfaction rating towards the AI Tutor in surveys before and after the certification program. The pre-certification surveys are distributed via and collected by Qualtrics when learners sign up to SDO. The post-certification surveys are distributed and collected on SDO immediately after learners complete learning activities (Supplementary Data 2).

We also evaluated the accuracy of AI Tutor’s answers to 298 bioinformatics questions using ten different LLMs. We compared the answers generated by the AI Tutor when it was powered by ten LLMs, including variants of Claude, GPT, Gemini, and LLaMA. For each model, we compared the AI-generated responses to a curated answer key reviewed by expert bioinformaticians. The AI Tutor-generated answers that differed from our vetted answer key were manually labeled as inaccurate. In June 2024, we ran this experiment 10 times using Claude 2^42^, Claude 3 Haiku^43^, Claude 3 Opus^44^, Claude 3 Sonnet^45^, Gemini 1.5 pro^46^, GPT-3.5^47^, GPT-4, GPT-4 Turbo^48^, GPT4o^49^, Llama2^50^ (Supplementary Data 3).

We also evaluated the effectiveness of AI Tutor’s guardrail by investigating the conversations where the AI Tutor told the learners it could not provide an answer (e.g. AI Tutor responded: I’m sorry, I don’t understand your question. Could you please ask a question related to bioinformatics, statistics, machine learning, artificial intelligence, Python programming, Jupyter notebook, R programming, genomics, epigenetics, transcriptomics, proteomics, microbiome, metabolomics, clinical data analysis, medical imaging, exposome, wearable data, cloud computing, or Pandas library?) We manually categorized the cases into four categories and calculated precision, recall, specificity, and F1 score (Supplementary Table 2 and Supplementary Data 4): (1) True Negatives (TN): The AI Tutor correctly identifies a non-SDO-content related question and refrains from answering it, as it is outside the specified domain of SDO-content; (2) True Positives (TP): The AI Tutor correctly identifies and appropriately responds to a SDO-content related question. This means the question is about SDO-content and the AI Tutor provides a correct and relevant answer; (3) False Positives (FP): The AI Tutor mistakenly identifies a non-SDO content related question as a relevant question, and attempts to provide an answer. This is an error as the AI Tutor is responding when it should not; and (4) False Negative (FN): The AI Tutor mistakenly identifies a SDO-content related question as non-SDO and fails to provide an answer, missing the opportunity to provide assistance to the students. This also includes answers that are incorrectly answered by the AI Tutor.

To understand the use cases of AI Tutor in learners’ learning experience, we evaluated 2,082 questions asked by the learners to the AI Tutor (Supplementary Data 4). We manually labeled the learning module topics (e.g., Programming) and the use cases of the AI Tutor corresponding to each learner-generated question (e.g., write or troubleshoot with code).

### Ethics and Data Governance

At the outset of the project, the research team consulted with institutional experts to ensure that systems and processes utilizing SDO reflected lifecycle data governance and oversight, especially data derived from human beings and sensitive categories of health data. Foremost among these were Stanford University’s IRB and Research Data Governance and Privacy Director based within the Vice Provost and Dean of Research. These authorities agreed that SDO’s systems and processes enable IRB oversight of data derived from human beings on a per-project basis. Studies that involve human subjects research or sensitive data must show that they have received IRB approval. The SDO platform itself was reviewed by the Stanford IRB and deemed non-human-subjects research. Therefore, learners’ informed consent is not required. In addition, the SDO’s anticipated use of data was reviewed and approved by Stanford University’s Privacy Office through its Data Risk Assessment process. All learners’ data are de-identified.

## Results

### Stanford Data Ocean Overview

SDO is designed to achieve three primary objectives: 1) robust data management that enables easy access to diverse multi-omics and wearable data, ranging from fully open access (e.g., COVID19 wearable datasets^39,52^) to partially restricted (Integrated Personal Omics Profiling project datasets^53,54^); 2) personalized education through the use of large-scale datasets and LLMs; and 3) cutting-edge research analytics that capitalizes on AI-driven visualization. Additionally, by transforming scientific papers into standalone learning modules—comprising datasets, code, and exercises—SDO accelerates research innovation while promoting reproducibility and collaborative knowledge sharing. To ensure sustainable learner engagement and long-term impact beyond certificate completion, SDO is creating a dynamic, interactive ecosystem that encourages continuous learning and professional growth. Our platform features monthly live seminars and workshops that not only keep learners updated on emerging technical skills but also foster the development of career-ready soft skills and professional networks. Moreover, learners are empowered to raise and discuss current challenges—such as the evolving interpretation of Variants of Unknown Significance (VUS) in genomic data—ensuring that they remain at the forefront of scientific advancements. Through initiatives such as collaboratively authored blog posts critiquing and replicating results of peer-reviewed research, sharing job opportunities on online discussion forums, and opportunities to collaborate on research projects, SDO provides a sustainable environment where learners can actively apply and expand their knowledge in real-world job and research contexts.

The platform achieves scalability by simplifying the initial setup and eliminating the need for extensive technical expertise and infrastructure maintenance. The platform also effectively enhances the learning experience through the integration of containerization and virtual machines, ensuring minimal interruptions during interactive engagement. The microservice architecture and real-time monitoring tools optimize performance and security, adhering to HIPAA standards. The platform also standardizes modules to promote consistency and reproducibility in bioinformatics research, supporting sustainable development in precision medicine. For more details, see Methods.

### Comprehensive Multi-Database Platform for Integrated Biomedical Data Analysis

SDO provides a multi-database platform designed to handle a wide variety of biomedical data types, including wearables, genomics, epigenomics, microbiome, metabolomics, and proteomics. This data diversity allows researchers to create comprehensive, real-time cohorts by integrating multiple data types, which can be pivotal for precision medicine research and personalized healthcare interventions. Users can seamlessly access and analyze these datasets through an integrated Jupyter notebook environment. The platform supports data from 107 iPOP subjects with 1416 visits in multiple longitudinal studies^53,54,55^ encompassing RNAseq, lipidomics, microbiome (gut 16s, nares 16s), metabolites, cytokine, targeted assays, and clinical test data (8637 datasets total). One hundred seven genome sequences are also available. Additionally, SDO includes publicly available tokenized data (heart rate, step, and sleep) for two COVID-19 studies: an early COVID-19 detection study of 5,300 participants (280 datasets and over 104 million data points)^52^ and a real time alerting study of 3,318 participants (4246 datasets and over 1.5 billion data points)^39^. These datasets enable SDO to offer extensive research and educational activities.

### Making Learning Precision Medicine Accessible

The curriculum, illustrated in Figures 1 and Supplementary Figure 2, outlines Fundamental Learning Modules covering foundational concepts such as Ethics, Programming, Statistics, Visualization, Cloud Computing, and Data Analysis (Multi-omics and Wearables), as well as Advanced Learning Modules that cover thematic areas such as Artificial Intelligence (AI) and Machine Learning (ML) techniques and applications in precision medicine. It integrates continuously updated content based on the latest research findings^56–59^ and includes interactive educational content using videos, guided Jupyter notebooks^60^, and exercises to support professional skill development. Supplementary Figure 2 shows a sample network curriculum on SDO structured around 24 Learn modules divided into six key thematic areas: Ethics, Programming, Statistics and Visualization, Cloud Computing, Data (which includes Multi-omics and Wearables), and AI/ML. Each module functions as an independent unit equipped with various educational artifacts such as videos, interactive notebooks, exams, self-evaluations, and practical exercises. This design is intended to cultivate a thorough understanding and proficient practical skills in each specific domain, ensuring that learners gain both theoretical knowledge and hands-on experience relevant to the field of bioinformatics and AI/ML.

**Figure 1.**
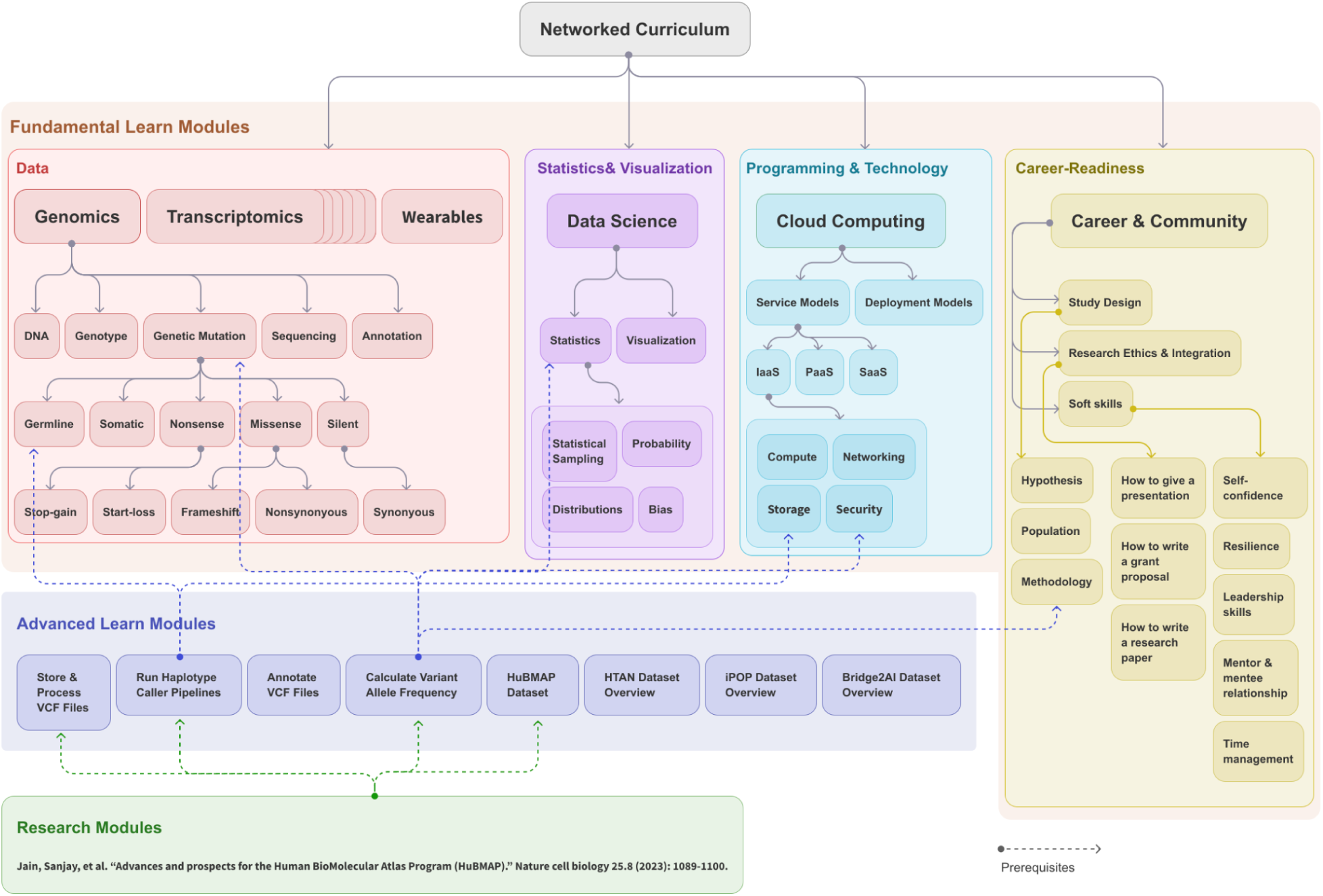
Comprehensive Networked Curriculum for Personalized Medicine Education on Stanford Data Ocean. A well-rounded curriculum in personalized medicine education includes networked modules in all relevant disciplines. This sample modular, networked curriculum consists of four major types of modules: 1) Fundamental Learn Module (grouped within a light peach-colored background): training material for understanding fundamental concepts around data (e.g., multi-omics), statistics and visualization (e.g., probabilities and distributions), programming and technology (e.g., cloud computing), and Career Readiness (e.g., communication, project management, and ethics). While each domain is visually differentiated with sub-color blocks, they are all part of the same light peach-colored foundational module group. Career Readiness includes a series of modules and live events designed to equip students with essential professional skills and industry insights, such as effective communication, teamwork, problem-solving, adaptability, and research capabilities including how to design a study, how to write a scholarly article, and how to present a paper. It also covers career-specific skills such as data privacy ethics, project management, and the use of AI tools in real-world scenarios. These modules aim to bridge the gap between academic learning and the demands of the workplace, preparing students for successful careers in bioinformatics and related fields; To ensure learners’ knowledge and skills stay up to date with industry demands in the long-term, we created new courses in AI/ML, and are building courses specialized in diabetes, cancer, and automating bioinformatics tasks with LLMs. The new courses are created in response to learners’ needs and advancements in AI-driven medicine, aiming to extend learners’ knowledge beyond the foundational modules into specialized niches. 2) Advanced Learn Module (purple background): a mix of multiple interdisciplinary concepts built on top of prerequisite fundamental modules (e.g., Processing large Variant Call Format (VCF) files needs the understanding of genetic mutation, basics statistics such as allele frequency, and how to store and process large files on the cloud); and 3) Research Module (green background): a combination of advanced and fundamental modules and provide more advanced information around a certain research topic (e.g., a research paper on Advances and prospects for the Human BioMolecular Atlas Program (HuBMAP) needs understanding of the HuBMAP dataset such as germline mutations and how to process large VCF files on the cloud in a secure fashion).

The platform supports individuals with varying educational backgrounds using a modular and networked curriculum, simplifying access without requiring software installations. This curriculum gradually introduces learners to bioinformatics, guiding them from basic concepts to advanced interdisciplinary topics such as biology, computer science, and statistics. This structured approach not only makes the field more accessible to beginners, but also accommodates personalized learning pathways, enhancing module reusability and keeping learners up-to-date with new developments^61^. Additionally, to help ensure accessibility to students of all technical levels and socio-economic backgrounds, SDO’s AI-driven tutor and visualization tools offer 24/7 assistance. The SDO’s modular course design enables educators to leverage a customizable curriculum by reusing existing modules and creating new ones. The train-the-trainer program provides educators with tools to implement instructional design best practices to effectively help students develop career-ready skills (see Career Readiness in Figure 1).

Based on the barriers reported by learners that made precision medicine education inaccessible for them (Supplementary Table 1), SDO offers low and middle income learners from all 50 states in the U.S. and 94 countries globally cost-free access to the Stanford Genetics certification in Bioinformatics and AI/ML. Low and middle income learners with free access make up 90.2% of SDO’s total learners population. Women make up 32.6%, projecting a positive long-term impact on the entire household’s education, healthcare, and income^62,63^. Post-certification, students are prepared to take responsibility for further study and continuous learning to sustain and grow their knowledge in the rapidly evolving field of continuous medicine.

### Learning Outcome and Satisfaction

We evaluated students’ learning outcomes and program satisfaction by measuring certificate completion rate, students’ self-efficacy, and perceived program impact.

The overall SDO Bioinformatics programs completion rate is 50.5%, and for structured cohort programs, it is 85.7%, both exceeding the Massive Open Online Courses (MOOCs) at 7%-10%^64–66^. In addition, certified students are required to achieve above 80% answer accuracy on formative assessment in quiz format following each learning module and 75% accuracy on the certificate exam following completion of all learning modules.

We measure self-efficacy by comparing students’ 1-5 Likert scale confidence ratings before and after each learning module on how much they agree they can achieve the module’s learning goals (see Supplementary Data 2). A significant growth in confidence ratings is observed across all learning modules, especially in Cloud Computing and Bioinformatics for learners from broad academic backgrounds (Supplementary Table 3 and 4). Among 1,495 learners, ratings of moderately confident and above increased by 33.9% to 97.6% for Cloud Computing and 47.8% to 96.1% for Bioinformatics (Figure 2). High academic self-efficacy in Science, Technology, Engineering, and Mathematics (STEM) is strongly associated with forming a science identity, taking more science courses, pursuing a science career, and predicting academic achievements^67–70^.

**Figure 2.**
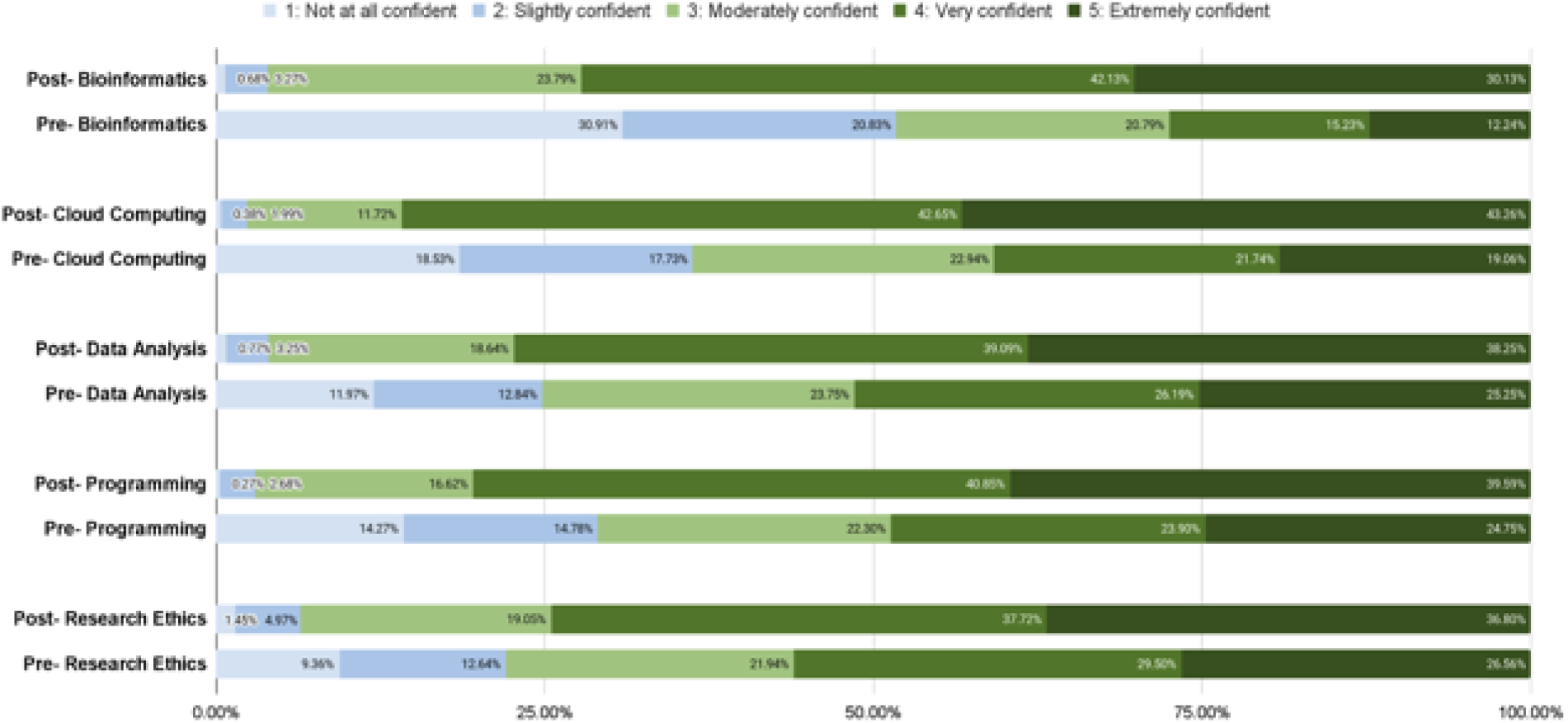
Enhanced Student Confidence in Precision Medicine through Bioinformatics Certificate Program. The confidence level on a sample of 1,495 learners on their competence regarding each learning topic were reported in surveys before and after they completed the Bioinformatics certificate program. The figure shows the noticeable increase in confidence in Cloud Computing and Bioinformatics topics (see Supplementary Table 3 and 4 for learners’ backgrounds). In the figure, light blue indicates the number of ratings that the learners are not confident at comprehending the key concepts in the learning modules; blue denotes the number of ratings that the learners are slightly confident at comprehending the key concepts in the learning modules; light green represents moderately confident; green represents very confident; dark green represents extremely confident.

Aligning with high self-efficacy’s career impact described in literature^67–70^, a survey of 72 certified respondents demonstrated high satisfaction. The program’s positive impact was reported at 97.5%, including 22.8% believing the programs positively impacted them securing internships or jobs, 27.8% developing interest to continue pursuing precision medicine, and 46.9% gaining confidence to apply for academic or professional opportunities. Additionally, we found 93.8% of certified learners wanted continued SDO involvement, either taking another course or teaching/ mentoring others. A total of 84.7% of certified learners recommended the program to others (Supplementary Data 1).

### AI Tutor

We built an LLMs-powered AI Tutor on SDO that democratizes private tutoring for students who cannot afford or allocate time for traditional methods, benefiting economically disadvantaged and underrepresented groups by providing an accessible and high-quality educational support. This AI Tutor (see Supplementary Figure 1b) specializes in questions pertinent to bioinformatics. It receives student inquiries, applies embedding techniques to identify the most relevant content within SDO, and uses prompt engineering to generate pertinent responses. In order to mitigate misinformation, the AI Tutor pulls answers from all the learning module content created and pre-vetted by Stanford scientists. For each AI Tutor generated answer, AI Tutor provides the source as SDO or LLM (see Supplementary Figure 1b). Learners can also clarify with or provide feedback to our teaching team during live office hours if they feel uncertain about the answers given by AI Tutor.

Every interaction is scrutinized under multiple layers of guardrails to ensure the accuracy of the information, prevent the generation of erroneous or misleading content (hallucinations), and maintain relevance to the field of bioinformatics. We have observed that hallucinations often occur when the system is asked to generate responses without sufficient context (e.g., The following question without the figure lacks context: Answer this question based on Figure 1 in Module 3.). In such cases, our guardrails prompt for clarification to ensure accurate and contextually appropriate answers. Similarly, if learners ask questions that lack context or fall outside the relevant bioinformatics domains, the AI Tutor will indicate that it does not understand the question and prompt learners to send another inquiries that are related to bioinformatics or its associated fields (e.g., statistics, machine learning, AI, genomics, etc.).

### Evaluating AI Tutor’s Performance

We evaluated the AI Tutor’s performance in three key areas: 1) Response Accuracy, based on its answers to 246 bioinformatics questions created by the SDO team; 2) Guardrails Performance, assessed through 2081 student-submitted questions; and 3) AI Tutor’s Use Cases and Perceived Usefulness.

### Response Accuracy

We compared 10 LLMs’ responses to 246 SDO-team-constructed bioinformatics multiple-choice questions with our answer keys. Figure 3 shows the performance of different LLMs: Claude 2^42^, Claude 3 Haiku^43^, Claude 3 Opus^44^, Claude 3 Sonnet^45^, Gemini 1.5 pro^46^, GPT-3.5^47^, GPT-4, GPT-4 Turbo^48^, GPT4o^49^, Llama2^50^. Initially, there were 298 questions, but 20 ambiguous questions flagged by the majority of the LLMs were removed.

**Figure 3:**
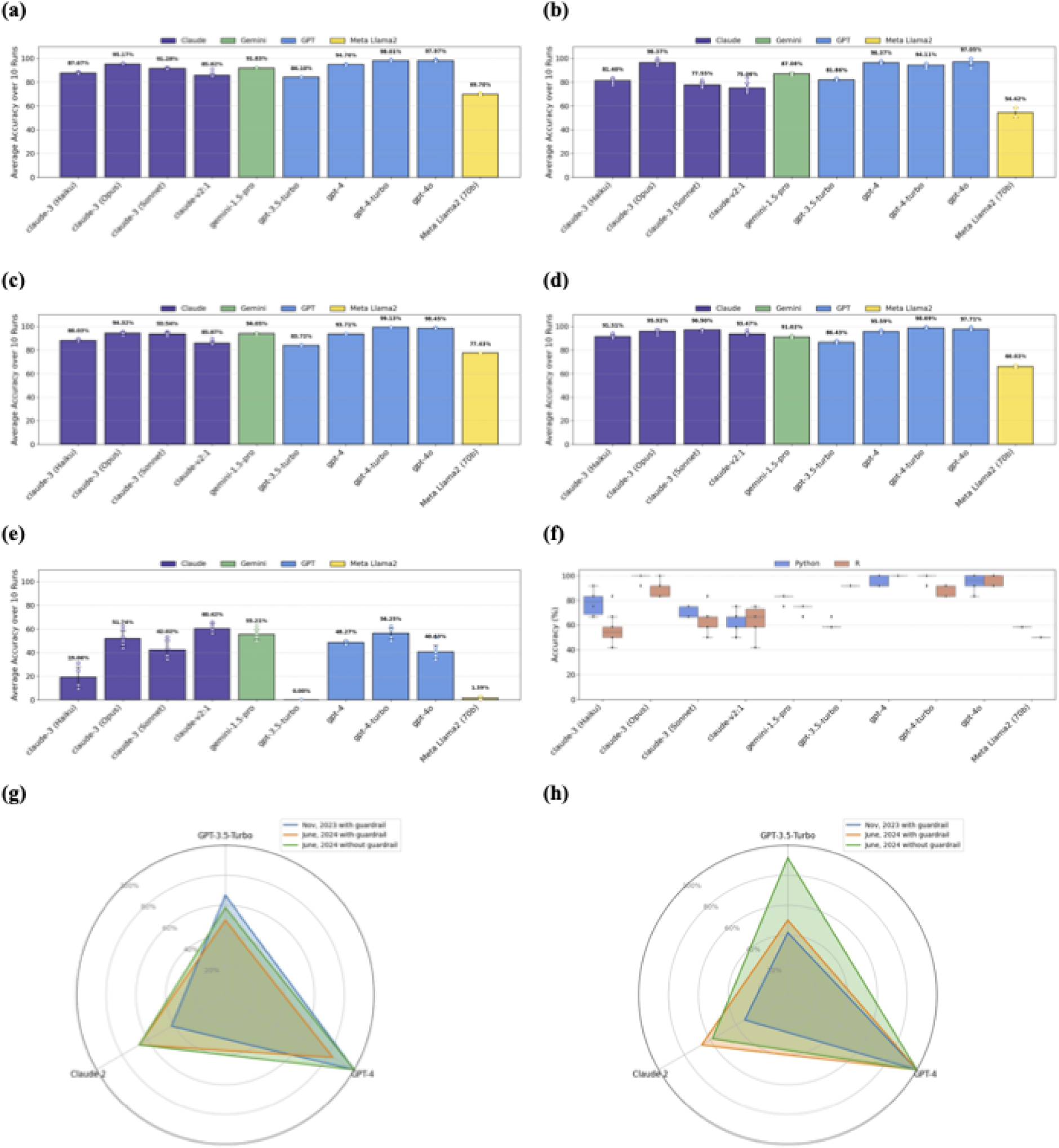
Performance of Different LLMs for 298 Bioinformatics Questions. **a**. Accuracy to General questions (n=246, excluding ambiguous and context-specific questions). This figure shows the accuracy of AI Tutor’s answers to 246 general questions after removing 20 ambiguous questions and 32 context-aware questions. Across Figure 3a-e, purple bars represent Claude models; the green bar represents Gemini model; blue bars indicate GPT models; the yellow bar denotes the Llama model **b**. Accuracy to Programming and Technology questions (n=49, excluding ambiguous and context-specific questions). Displays the performance of LLMs for programming (R, Python, Pandas) and cloud computing questions (49 questions). **c.** Accuracy to Statistics and Visualization questions (n=129, excluding ambiguous and context-specific questions). This panel illustrates the performance of LLMs for statistics and visualization questions (129 questions). **d.** Accuracy to Research Ethics, Multi-omics, and Wearable Data (n=68, excluding ambiguous and context-specific questions). This panel depicts the performance of LLMs in answering questions related to research ethics, multi-omics, and wearable data questions (68 questions). **e.** Accuracy to context-specific questions (n=32). This panel presents the performance of LLMs for context-aware questions (32 questions); the model was expected to either request additional context or indicate that it lacked the necessary context to respond accurately. Responses that failed to acknowledge the need for context were considered incorrect. **f.** Accuracy to questions about Python (n=12) vs. R (n=12). comparing the performance of different models for Python (12 questions) vs. R (12 questions), showing some models are biased toward Python and cannot identify the R context well. In this figure, blue represents Python, and red represents R. Bars show mean accuracy across n = 10 independent runs per condition; error bars represent ± 1 SD. g. Python (n=12) (Nov. 2023 vs. June 2024) and h. R (n=12) (Nov. 2023 vs. June 2024). Panels (**g**) and (**h**) compare performance of GPT-3.5-turbo, GPT-4, and Claude 2 for Python (12 questions) and R (12 questions): November 2023 vs. June 2024 (with and without guardrail). Unlike in Figure 3a-f, where we used the bare LLM (Execution Engine), for G and H, we employed the AI Chatbot (see Supplementary Figure 1b). Both the Execution Engine and the Guardrail utilized the same LLM. There are variations in performance in how these models differentiate between Python and R coding questions. Claude 2 showed significant improvement in handling R/Python questions. Similar findings are reported in recent studies51. Another observation concerns guardrails: GPT-3.5 flagged 5 out of 12 Python questions and 4 out of 12 R questions as irrelevant. It is important to consider while designing guardrails—none of the questions were flagged by GPT-4 as irrelevant in 2023 or 2024. The closed-source nature of some LLMs raises concerns about predictability and interpretability, particularly in medicine, where decision-making is paramount. For both Figures 3g-h, blue represents the AI Tutor we used in November 2023 with guardrail; orange represents the AI Tutor we used in June 2024 with guardrail; green represents the AI Tutor we used in June 2024 without guardrail for this particular experiment while AI tutor in production is equipped by guardrails.

In our study, we grouped general questions into three categories as depicted in Figure 1: Programming and Technology; Statistics (including Visualization and AI/ML); and Data (covering Multi-omics and Wearables). We conducted a detailed analysis of various LLMs’ performances across these categories, as shown in Figure 3a-d, evaluating them on general and domain-specific multiple-choice questions relevant to bioinformatics topics for 10 times in June 2024 (Supplementary Data 3). To evaluate the risk of LLMs giving erroneous answers without sufficient context, we compared the LLMs’ accuracy of 32 questions (i.e., context-aware questions) that refer to specific information inaccessible to the LLMs, such as an image, a code block, or a research study (Figure 3e). In healthcare applications, when a model attempts to interpret missing data that a physician failed to provide without acknowledging it (i.e., hallucinating), the LLM-generated information could lead to significant health and financial cost. We also examined the models’ differential responses to Python and R programming questions, noting a tendency in some LLMs to favor Python, which often leads to mistakes in R contexts (Figure 3f). This bias was further analyzed in Figures 3g and 3h, comparing the performance of models such as GPT-4, GPT-3.5-turbo, and Claude 2 over two periods, November 2023 and June 2024.

The findings from our figures indicate that overall, the GPT-4 family generally outperforms other models across most question categories (Figure 3a-d). GPT-4o achieved the highest accuracy in General questions as well as Programming and Technology questions (Figure 3a and 3b), whereas GPT-4-turbo excelled in Statistics and Visualization questions, as well as Research Ethics, Multi-omics, and Wearable Data questions (Figure 3c and 3d). The Claude 3 family also performed strongly, particularly Claude 3 (Opus), which shows high accuracy in multiple categories. However, GPT-3.5-turbo performed poorly on context-specific questions (Figure 3e), indicating significant limitations in handling queries that require specific contextual understanding. The comparison of Python vs. R responses (Figure 3f) reveals a notable bias towards Python, with some models performing significantly better in Python than in R. Furthermore, the radar charts (Figure 3g and 3h) illustrate the improvements or regressions in model performance over time, with Claude 2 showing notable improvements in handling guardrail scenarios in June 2024 compared to November 2023.

### Evaluating AI Tutor’s Guardrail Performance

We implemented the AI Tutor using GPT-4 in production as the default LLM, although learners can switch to other LLMs via the user interface. To ensure the AI Tutor responds only to questions related to the educational content in SDO, we built several guardrails (see Supplementary Figure 1b). Given that LLMs are constantly evolving, which could result in unpredictable AI Tutor responses, we set constrained guardrails to mitigate risks while ensuring the AI Tutor can effectively support learning activities.

The performance of the 2,082 AI Tutor responses to students’ questions demonstrated its ability to adhere to the guardrails (Supplementary Table 2). The guardrail’s precision was calculated at 100%, indicating no false positives among the predicted positives. Recall, or sensitivity, was 93.4%, reflecting that most positive cases were correctly identified. Specificity was 100%, meaning all true negatives were accurately recognized. Additionally, the F1 score, the harmonic mean of precision and recall, was 96.6%, providing a balanced measure of the model’s accuracy in identifying both classes. Notably, there were 126 false negatives, indicating missed SDO-content-related questions. These metrics indicate AI Tutor is capable of providing correct and relevant answers to students, cultivating trustworthy interactions with students.

### Evaluating AI Tutor’s Use Cases and Perceived Usefulness

SDO’s blended learning model effectively combines self-paced modules with interactive AI-facilitated learning activities, enhancing educational engagement and outcomes. Students utilizing this hybrid approach not only benefit from the flexibility of independent study but also gain substantial support through structured digital interactions, as shown by multiple studies^71,72^. This methodology particularly benefits underprivileged students, offering frequent opportunities to interact with AI tools, which is critical for developing AI literacy. LLM applications have demonstrated creativity surpassing 99% of people in originality and fluency. GitHub Copilot sped up coding tasks by 55.8%. Proficiency in AI tools not only boosts learners’ project and career prospects in precision medicine but also enables learners to become efficient collaborators in research and reduces training periods in professional settings^73–75^.

Figure 4a shows that 46.7% of the most queried topics include Programming and Cloud Computing, while 21.2% cover Statistics, Visualization, AI/ML, and 20.3% relate to Bioinformatics/Omics-data. As for the types of questions, Figure 4b indicates that 39.5% of the inquiries involve clarifying or troubleshooting code, 18.2% ask about a quiz or exam, 13.8% are statistics questions, and 11.8% are bioinformatics questions.

**Figure 4.**
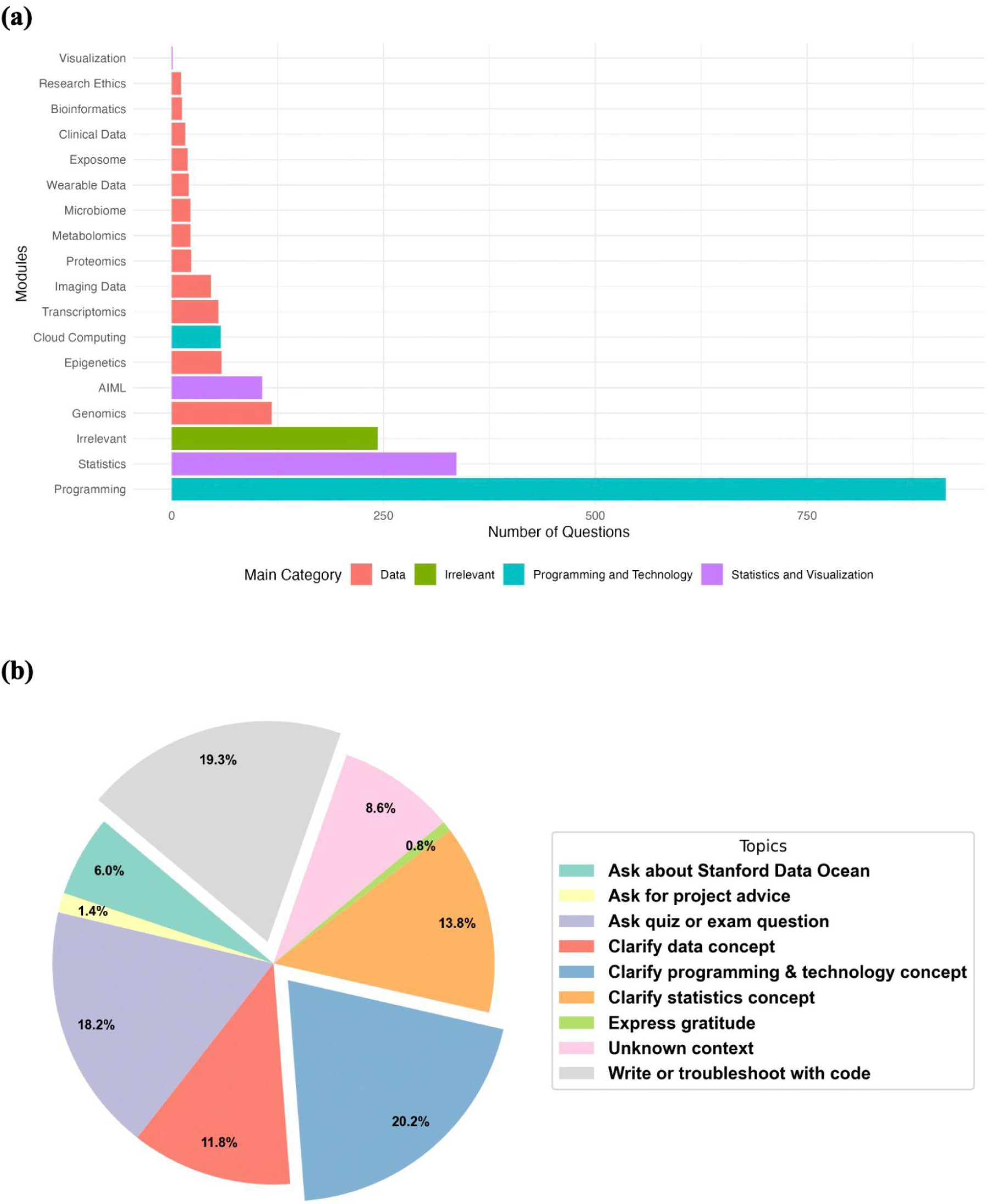
Learner Engagement Analysis with AI Tutor. **a**. Learners’ Questions Categorized by Learning Modules Topics (n=2,082). A total of 2,082 questions from 156 learners categorized by learning modules topics (Supplementary Data 4). 43.9% of the questions are about Programmings, followed by 16.1% Statistics. a(ii) 47.3% of the questions labeled as Irrelevant are questions about Stanford Data Ocean. 19.3% are questions that lack context to identify students’ intentions.In this figure, red color in the figure indicates learner questions related to omics data or wearable data; green denotes learner-generated queries that are irrelevant to bioinformatics (e.g., greetings, expressing gratitude); blue corresponds to questions relevant to Python, R, and Cloud Computing; purple pertaining to to statistics and data visualization. **b**. Learners’ Questions Categorized by Goals (n=2082). A total of 2,082 questions are categorized by the goal students are trying to accomplish by using AI Tutor. 20.2% are about clarifying programming and technology concepts, and 19.3% are about writing or troubleshooting code. We gave the AI Tutor a persona and described what it is and what it can do (e.g., The prompt: You are a helpful assistant well-versed in bioinformatics and related technologies. Please answer questions with all the needed context related to the Fundamental Modules content multi-omics, statistics, R, Python, etc. If a question pertains to a different topic, politely refuse to answer.) In this figure, turquoise color represents learner-generated questions to the AI Tutor about the SDO platform’s functionalities and user experience; yellow denotes specific questions about learner’s individual projects; purple represents questions related to the SDO’s quiz or exams; red represents questions that are trying to clarify unfamiliar concepts in omics data or wearable data; blue represents questions that are trying to clarify unfamiliar concepts in Python, R, or Cloud Computing; orange represents questions that are trying to clarify unfamiliar concepts in statistics or data visualization; green represents queries that expresses thankfulness to the AI Tutor’s answers; pink marks queries that lacks context; grey indicates questions that ask the AI Tutor to write or explain the code or help them troubleshoot the code that generated errors.

A quantitative evaluation of the AI Tutor’s impact shows high student satisfaction and perceived effectiveness. On the platform, 76.56% of responses were positively rated, and 71.3% of students on a six-point Likert scale strongly agreed that the AI Tutor enhanced their understanding of precision medicine (see Supplementary Data 2). Additionally, 21.3% moderately agreed, and only 6.4% slightly agreed. The AI Tutor excelled in programming and bioinformatics—key areas of precision medicine—with particularly high agreement in technology-related modules such as statistics and cloud computing. These results highlight the AI Tutor’s effectiveness in supporting student learning and identify potential areas for further improvement to optimize user experience.

### LLM-based Research Data Visualization

#### AI Tutor for Data Visualization: Fostering Algorithmic Thinking

Analysis of learner queries in Figure 4b reveals that 39.5% of learners concentrate their questions on aspects of programming, such as interpreting and troubleshooting code. This observation prompts a new research question emerging directly from the data: Is extensive programming knowledge essential for completing precision medicine tasks? The frequency of programming-related inquiries and the effectiveness of the AI Tutor by learners underscores the need to reevaluate and enhance how educational models integrate programming skills with domain-specific scientific training.

Programming fosters critical thinking and problem-solving skills^76^ (e.g., through Divide-and-Conquer, Dynamic Programming, Greedy Algorithms, Graph Algorithms, Probabilistic and Analysis, and Randomized Algorithms). However, while error handling builds resilience, persistence, and adaptability, it is time-consuming and detracts from valuable research hours that could be used to unlock biological mechanisms. To address this, SDO’s AI Data Visualization tool prioritizes algorithmic thinking using LLMs—shifting the focus away from memorizing code syntax and time-consuming debugging. This approach helps learners in the life sciences develop higher-order thinking skills, particularly in problem-solving and critical reasoning^77^.

The SDO’s AI Data Visualization tool enables users to import their own biomedical datasets for research analysis. This tool supports multi-modal analysis, accommodates a broader array of data formats, and incorporates automatic error handling, all while being compatible with both Python and R. For more details on this innovative approach, see Methods. The visualization component operates under the assumption of two primary user groups:

1. Non-technical Users: Users with no programming experience or data familiarity use system generated prompts. Here, the platform first summarizes the dataset and leverages LLMs to generate an explanation of the data and potential visualization goals (i.e., prompts); users can then select their desired goal, prompting the system to utilize the LLM output and summary to automatically generate visualization code and produce the corresponding plots. This multi-step process (e.g., data summarization, goal generation, and elaboration, reading user-generated prompt, code creation, error handling, and plot generation) is further detailed in Supplementary Figure 1c. Users can further explore the data by posing questions, with the platform assisting in code generation and visualization based on the provided query. Figure 5 illustrates the capability of SDO AI-facilitated visualization for autonomous interpretation and visualization of multi-omics and wearable sensor data (Supplementary Data 5).
2. Technical Users: Users with an understanding of data analysis but want to save time from low-level programming challenges such as syntax, library management, debugging, and API updates (e.g., physicians, geneticists, or biologists). These users simply input the desired algorithm (see Supplementary Figure 1c), prompting the system to generate the code and corresponding visualization. It is crucial to note that problem definition and algorithmic thinking—defined as the ability to break down problems into a series of logical steps—are essential elements of this process. Proper problem definition and algorithmic thinking are vital because they guide the AI in generating accurate and relevant code; without these, users may encounter incorrect or inefficient solutions. The AI Tutor supports users by helping them better define their problems and develop algorithmic thinking skills, ensuring the system produces the most appropriate code and visualizations. We showcase the versatility of the SDO platform in addressing various research questions posed by technical users (Figure 6). Figures 6a-6e demonstrate the robustness and versatility of automated code generation for replicating and interpreting complex visualizations across diverse research domains (Supplementary Data 5).

**Figure 5:**
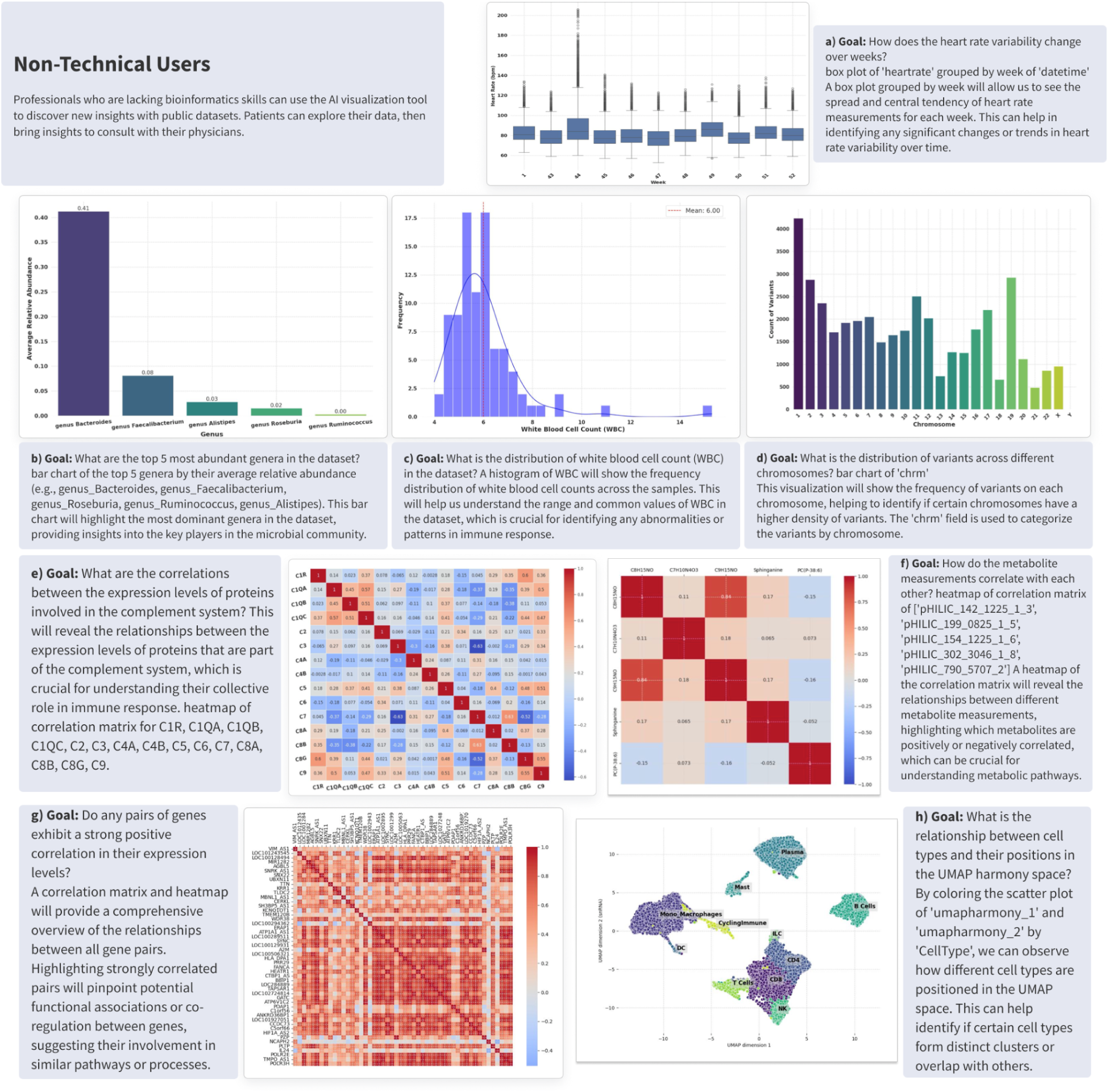
Autonomous Interpretation and Visualization of Multi-Omics and Wearable Data through Language Model Integration. The text boxes represent goals expressed as prompts, generated by GPT-4o based on summary data and metadata from the SDO platform. **a.** Heart Rate Variability Over Time (Wearable data): Box plot showing weekly heart rate variability across different weeks, providing insights into changes in heart rate dynamics over time. **b.** Dominant Bacterial Genera (Gut 16S data): Bar graph illustrating the relative abundance of the top five most abundant bacterial genera in the dataset, highlighting key players in the microbial community. **c.** White Blood Cell Count Distribution (Clinical data): Histogram showing the frequency distribution of white blood cell counts, crucial for identifying abnormal immune response patterns. **d.** Chromosomal Variant Distribution (Genomics data): Bar chart representing the frequency distribution of genetic variants across different chromosomes, useful for identifying potential chromosomal hotspots of variability. **e.** Protein Interaction in Immune Response (Proteomics data): Correlation matrix displaying relationships among protein expression levels involved in the complement system, aiding in understanding their collective roles in immune response. **f.** Metabolite Intensity Correlations (Metabolomics data): Heatmap showing correlations between spectral intensity measurements of metabolites, revealing interactions and dependencies crucial for metabolic studies. **g.** Gene Expression Correlations (Transcriptomics data): Correlation matrix and heatmap analyzing pairwise relationships between gene expression levels across the genome, providing insights into potential regulatory and co-regulatory networks. H. Immune Cell Distribution (snRNA data): UMAP representation of snRNA immune cells colored by cell type, illustrating the relationship between cell types and their positions in the UMAP harmony space. Users can fine-tune the visualization by providing feedback, such as changing the x-axis title. The data used in this demonstration were sourced from four independent studies: Miss^52^ (Participant ID: A0NVTRV for Figure 5a), Zhou et al.^53^ (Participant ID: ZOZOW1T for Figures 5b, 5-c, 5e, 5f, 5g), the 1000 Genomes Project^78^ annotated by the COSMIC68 dataset^79^ for Figure 5d, and snRNA immune cells processed data from Hickey et al.^80^.

**Figure 6:**
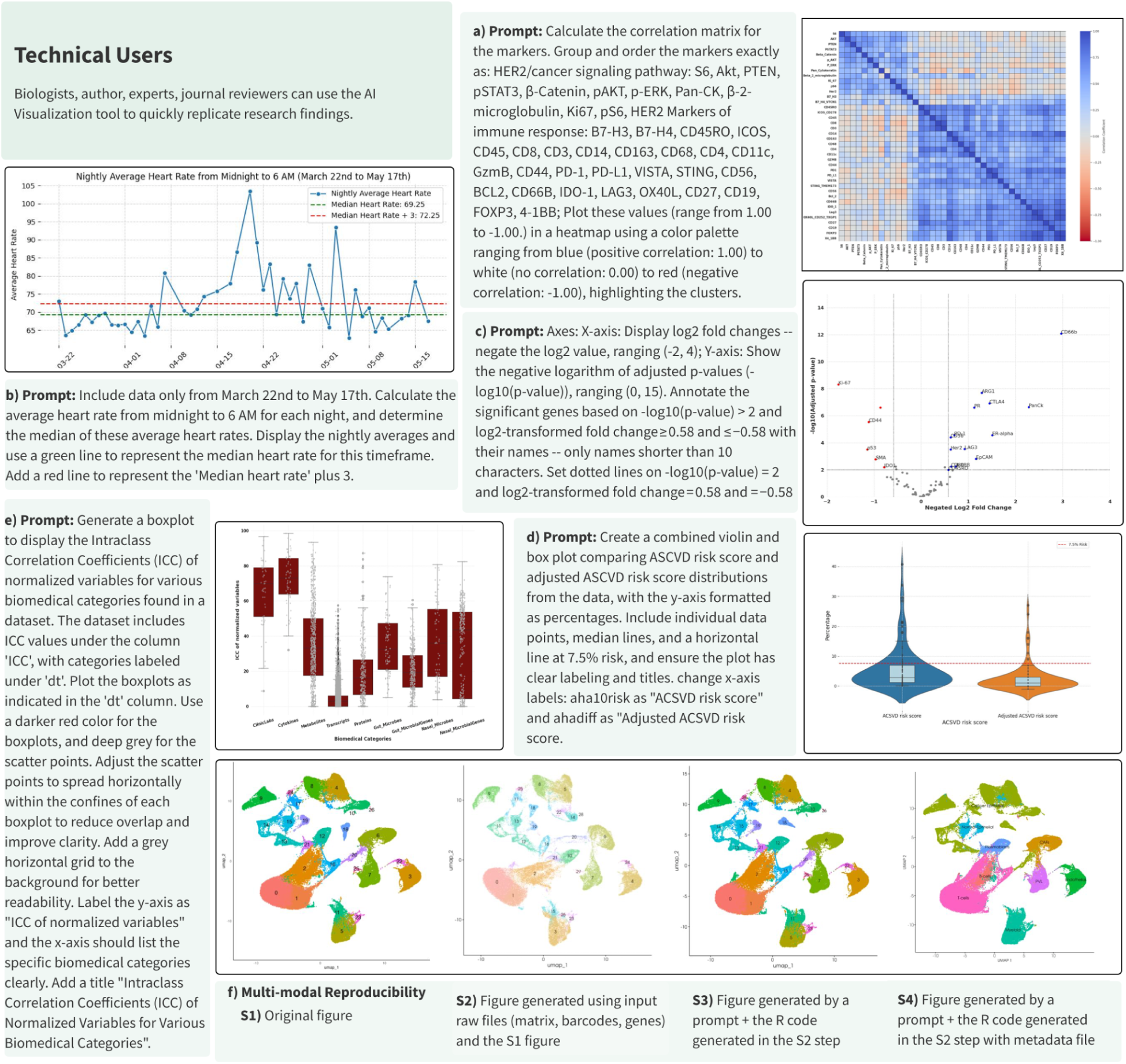
Automated Code Generation and Execution from Natural Language Inputs for Data Visualization in Multi-Omics and Wearables Research. The text boxes represent expert-curated prompts designed to reproduce each figure using public multi-omics and wearable datasets. **a.** Protein Marker Correlation Analysis: Heatmap displaying pairwise correlation coefficients for protein markers involved in pathological complete response (pCR) analysis. This figure replicates and expands upon the heatmap from Figure 2.c in McNamara et al.^81^, highlighting potential biomarker interactions in cancer pathways. **b**. Heart Rate Analysis for COVID-19 Detection: Line graph depicting daily average resting heart rates from March 22 to May 17th, identifying significant trends that could indicate physiological responses to COVID-19. This visualization is inspired by the NightSignal algorithm and replicates the scenario depicted in Figure 1.b in Alavi et al.^39^, illustrating variations in resting heart rate during the pandemic. **c**. Differential Protein Expression in Oral Cancer: Volcano plot contrasting protein expression profiles between bacteria-positive and -negative regions in oral squamous cell carcinoma. Adapted from Figure 2 in Galeano Niño et al.^82^, this plot aids in understanding micro-niche level variations within tumor environments. **d.** ASCVD Risk Score Distribution Analysis: Combined violin and box plot of atherosclerotic cardiovascular disease (ASCVD) risk scores, comparing standard and adjusted scores across a sample. Based on Figure 4a in Schössler-Fiorenza Rose et al.^54^, it facilitates risk assessment and stratification in clinical research. **e**. Variance Analysis in Multi-Omics Data: Boxplot showing the intra-class correlation coefficients for various biomedical categories, indicating variance levels attributed to participant structure. This visualization is based on Figure 2a in Zhou et al.^53^, which is crucial for evaluating consistency across multi-omic datasets. **f**. scRNA-seq Data Visualization via UMAP (Uniform Manifold Approximation and Projection): UMAP plot derived from scRNA-seq data files (barcodes.tsv, genes.tsv, matrix.mtx), visualizing gene expression patterns across different cell types. The AI visualization component summarizes all input files and iteratively generates the plot. While the output UMAP in Fig F.S2 is not exactly the same as F.S1, by adding the right set of steps, the SDO AI visualization component successfully captured the clusters in F.S2. The prompt is described as the following: Filter the cells in the Seurat object to include only those with an RNA count of less than 100,000. Normalize the data in the Seurat object using the LogNormalize method, with a scale factor of 10,000. Identify the top 2,000 variable features (genes) in the dataset using the variance-stabilizing transformation (VST) method. F.S4 was built on top of F.S2 and a metadata file. The pipeline in each step provides code and documentation of the thought process it went through to generate the visual. The user can use the documentation to guide the model, where necessary, to achieve the desired outcome (image). This process is iterative.

Reproducibility Feature: Reproducing plots can be challenging due to authors potentially neglecting key parameters such as unclear naming conventions, data/plot inconsistencies, and inadequate data type specifications. Incomplete or unclear documentation on how to install and run code can pose a significant challenge to replication, especially for researchers who may not be well-versed in the necessary tools and package managers^83,84^. These inconsistencies significantly complicate reproducibility through conventional programming methods. To address this challenge, we have integrated an additional feature within the toolset. Users can provide the target figure and corresponding dataset, prompting the system to leverage LLMs and error handling to reproduce the plot. While successful in many cases, instances of author-specific assumptions (unclear and inconsistent documentation) have rendered figure reproduction nearly impossible despite repeated LLM attempts. Consequently, users must intervene and provide feedback to the LLMs, nudging them to address these issues.

Fig. 6f represents an application of single-cell RNA sequencing data visualization. It employs a UMAP technique to visualize data clusters based on the raw files from Wu et al.^85^. Despite inherent variations due to different computational tools and the stochastic nature of UMAP, SDO effectively captures and displays the main clusters, highlighting its adaptability and accuracy in handling complex genomic data. The platform handles new data types and complex multimodal visualizations.

We propose a potential solution to enhance reproducibility: encouraging authors to provide prompts for future plot generation. This practice could incentivize the explicit communication of implicit assumptions during the visualization process, improving reproducibility.

## Discussion

Our results demonstrate that a serverless platform leveraging cloud computing can substantially address education challenges in bioinformatics, providing improved access to data and computational resources, particularly for economically disadvantaged and historically marginalized populations. The increase in learners’ competence in bioinformatics skills (Figure 2) and professional advancement mirrors previous findings that LLM tools improve students’ learning performance and higher-order thinking^86^. Furthermore, we recognize the challenges associated with extensive time spent on debugging and synchronization for non-programmers in our analysis on AI Tutor chat logs (Figure 4a and 4b), alongside the lengthy upskilling/reskilling required for scientific professionals with non-programming trainings (Supplementary Table 1). Aligning with previous research^75^, our platform demonstrates how LLM-enabled solutions similar to SDO can reduce programming barriers to boost team productivity. The capabilities of the AI Data Visualization tool demonstrated in Figure 5 and 6 enable professionals, regardless of their technical backgrounds, to leverage and improve algorithm thinking to tackle the computational aspects of complex bioinformatics problems. AI-powered educational tools similar to SDO help learners focus on how to approach and solve real bioinformatics problems, instead of getting stuck on coding details. This makes it easier for people from different backgrounds to understand and apply key ideas in data analysis.

The emergence of LLMs presents a new set of challenges. The potential for plagiarism through copy-pasting generated content using LLMs necessitates the development of robust anti-cheating measures (Figure 4b). While solutions such as CodeHelp^87^ incorporate guardrails designed to prevent the direct revelation of solutions, thereby aiding students in resolving their issues ethically, the challenge remains that students may still access LLMs lacking guardrails. We hypothesize that incorporating context-specific questions could further mitigate the risk of cheating by reducing the accessibility of inappropriate assistance. Our analysis of the AI Tutor’s responses (Figure 3) show the guardrail is effective in identifying questions that are relevant for bioinformatics related content, and provide accurate and relevant responses. This approach ensures that the AI support remains aligned with educational goals and maintains academic integrity. Furthermore, standardization and reproducibility issues persist^83,84^, impeding the achievement of consistent, reliable results essential for validating findings and ensuring trustworthy scientific advancements. One potential solution involves encouraging authors to create, test, and submit their prompts alongside published manuscripts.

We found that answer accuracy improved across three successive releases of commercial models (Claude, GPT, and Gemini in Figure 3), whereas Llama 2-70B we tested showed comparatively lower accuracy. Since we evaluated only one open-source model at a single time point, we recommend that future precision-medicine benchmarks include multiple open-source releases to determine whether freely available models can reach the accuracy thresholds necessary to develop effective LLM-enabled tools for precision medicine research and education, particularly in under-funded institutions and economically disadvantaged communities^88^.

## Code Availability

The software platform supporting the findings of this study is available at https://dataocean.stanford.edu/. Access to the platform can be granted upon request. Interested parties should visit the website to initiate a request for platform access.

## Data Availability

The program’s impact is from Supplementary Data 1. The source data for Figure 2 can be accessed from Supplementary Data 2. The source data for Figure 3 can be accessed from Supplementary Data 3. The source data for Figure 4 and Supplementary Table 2 can be accessed from Supplementary Data 4. The prompts for Figure 5 and Figure 6 can be accessed from Supplementary Data 5. The source data for Supplementary Table 3 and 4 can be accessed from Supplementary Data 6. The datasets supporting the findings of Figures 5 and 6 are publicly accessible, as detailed in the Data Availability sections of the respective source papers^39, 52, 53, 54, 78, 79–82^. For the Stanford Data Ocean’s (SDO) learners’ pre- and post-surveys, as well as the AI Tutor questions, all personally identifiable information has been removed to ensure privacy and confidentiality. This includes the deletion of email addresses, first and last names, and any other information that could be used to identify individual participants.

## Data Availability

The datasets supporting the findings of Figures 5 and 6 are publicly accessible, as detailed in
the "Data Availability" sections of the respective source papers. For the Stanford Data Ocean's
(SDO) learners' pre- and post-surveys, as well as the AI Tutor questions, all personally
identifiable information has been removed to ensure privacy and confidentiality. This includes
the deletion of email addresses, first and last names, and any other information that could be
used to identify individual participants.

## Acknowledgments

We acknowledge Amazon Web Services, Microsoft Azure and OpenAI for this research. This research also received support from Stanford Institute for Human-Centered Artificial Intelligence (HAI) and Stanford Accelerator for Learning. We acknowledge the Stanford Genetics Bioinformatics Service Center for providing this research’s gateway to the SCG cluster, Google Cloud Platform, and Amazon Web Services. We would like to extend our special thanks to Steven Gin, Heather Matson, Claudiu Farcas, Andrew Crabb, Sujaya Srinivasan and Carina Kemp from AWS for their invaluable guidance and support throughout this research. We gratefully acknowledge Conectado Inc. for their crucial support in the outreach and recruitment of underserved students, with special thanks to Guillermo Diaz Jr. and Diana Pacheco for their partnership and dedication. We also thank Fred Swaniker from ALX Africa and Sand Technologies for their support in expanding access to this research and learning initiative.

## Contributions

Study conception, platform design & build: A.B., K.C., A.A., A.D. Manuscript drafting: A.B., K.C., A.A., A.D.M.; Experimental design & data analysis: A.B., K.C., A.A., A.R., S.M., R.A.S. Learning & Research-module development and evaluation: A.B., K.C., A.A., A.R., R.P., F.G., R.N., X.Z., M.W., J.L.P.T., I.A., J.Y., A.T., S.M.S.-F.R., M.B., K.Y., X.Zh., D.J.F.R., A.K., J.W.; Infrastructure architecture: A.B., P.S., A.D., A.A. Structured workshop organisation & delivery: A.B., K.C., A.R., E.M., J.L., J.Y., K.D., M.S. Ethics review & oversight: E.M.M., S.E., C.N., J.Y. Project supervision & funding acquisition: A.B.; All authors reviewed and approved the final version.

---

**Supplementary Figure 1:**
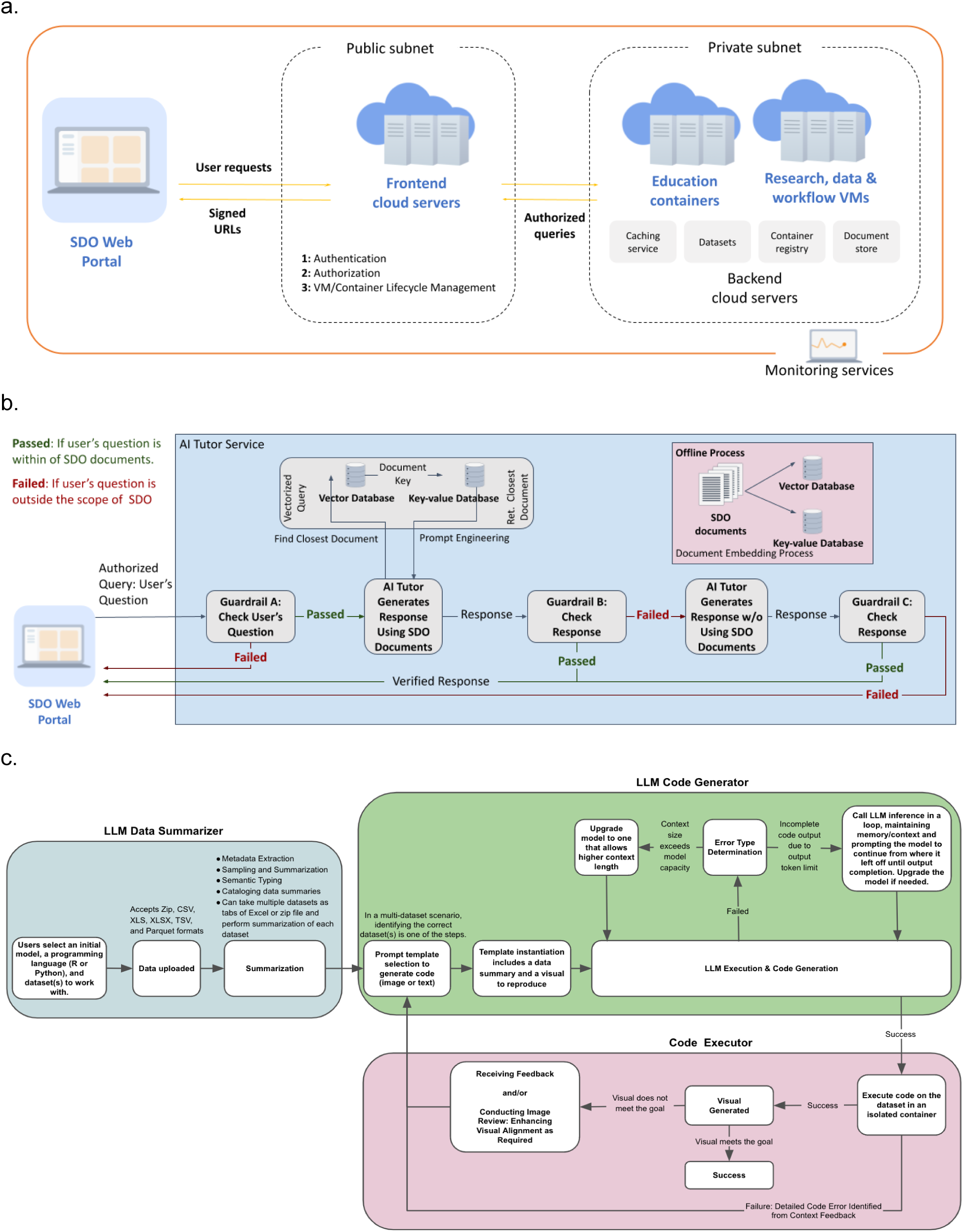
Stanford Data Ocean Architecture. (a) Architecture of the Platform (Data, VMs/Containers, and Workflows) (b) AI Tutor as a Chatbot (c) AI Tutor for Data Visualization

**Supplementary Figure 2:**
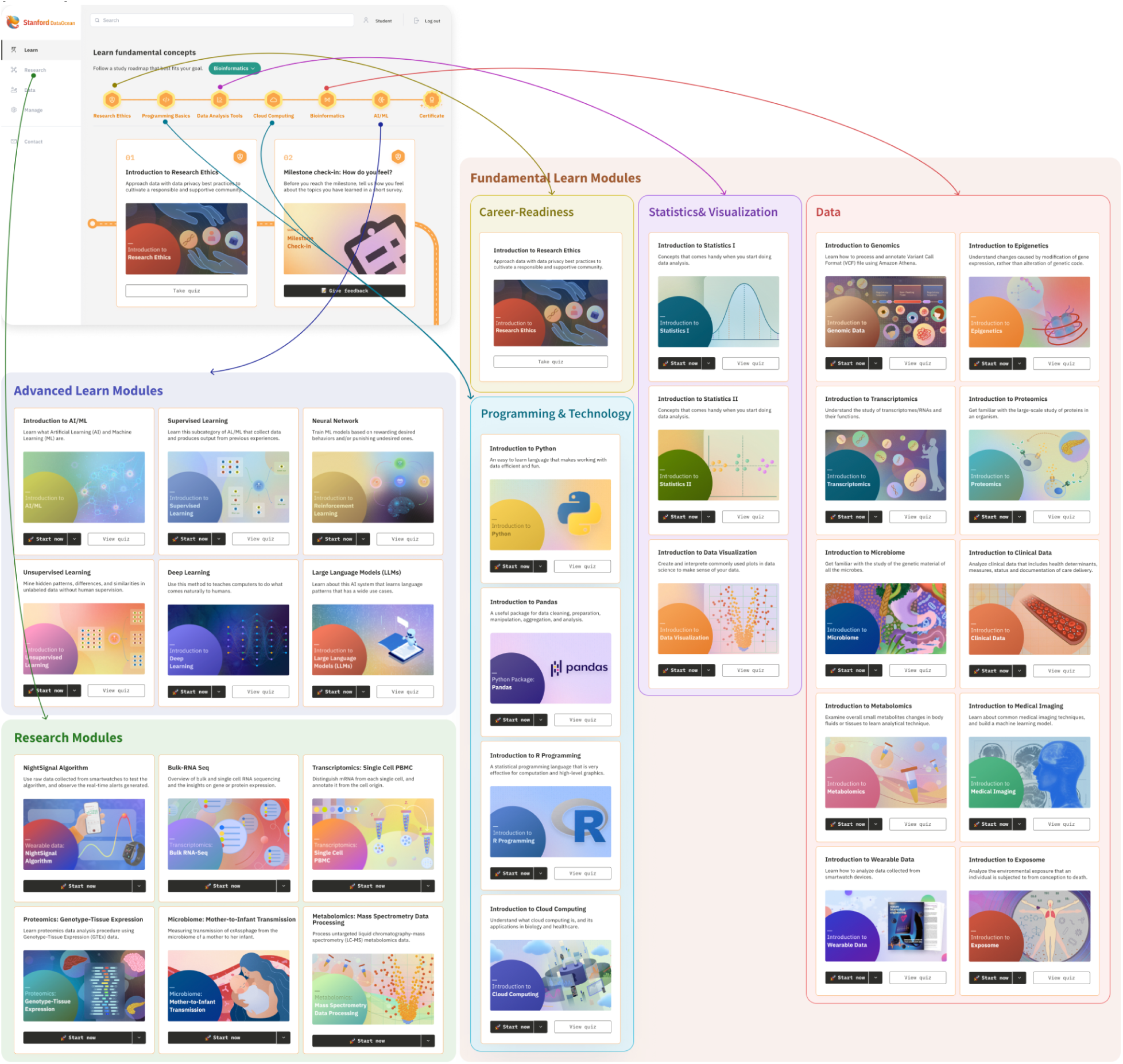
The integrated network curriculum on the Stanford Data Ocean (SDO)

**Supplementary Table 1.** Summary of Challenges Faced by a Sample of 381 Scholarship Applicants (June 2024)

| Challenge Category | Application Percentage | Application Quantity | Affected Demographics/Regions |
| --- | --- | --- | --- |
| Financial Constraints | 73% | 278 | Low-income families, immigrants, international students residing in developed nations, and residents in countries facing economic crises or sanctions (e.g., Nigeria, Turkey, Iran) |
| Lack of Interdisciplinary Medical Education | 20.1% | 80 | Residents in nations with early Precision Medicine development, rural areas, or politically unstable regions (e.g., Ukraine) |
| Limited Time Due to Responsibilities | 6% | 23 | PhD/MD students with busy schedules, individuals with multiple jobs or caring duties |

**Supplementary Table 2.** AI Tutor Performance Metrics (2,082 AI Tutor’s responses to 2,082 learner-generated questions) (Supplementary Data 4)

| Category | Definition | Value |
| --- | --- | --- |
| True Negatives (TN) | The AI Tutor correctly identifies and ignores questions that are not related to SDO content. | 170 |
| True Positives (TP) | The AI Tutor accurately identifies and appropriately responds to SDO-content-related questions. | 1,786 |
| False Positives (FP) | The AI Tutor incorrectly responds to non-SDO-content-related questions. | 0 |
| False Negatives (FN) | The AI Tutor fails to recognize or incorrectly answers SDO-content-related questions. | 126 |

**Supplementary Table 3.** Prior Experience of a Sample of 1,495 Graduates from Bioinformatics Program (Supplementary Data 6)

| Prior Experience | Percentage |
| --- | --- |
| Computer Science | 20.9% |
| Biomedical Data Science/ Bioinformatics | 9.1% |
| Other | 12.5% |
| Mathematics | 8.9% |
| Medicine | 8.2% |
| Computer Engineering | 8.4% |
| Statistics | 9.5% |
| Physics | 4.4% |
| Social Sciences | 3.7% |
| Business | 5.3% |
| Electrical Engineering | 2.7% |
| Art & Humanities | 2.5% |
| Biomedical Engineering | 2.5% |
| Biochemical Engineering | 1.5% |

**Supplementary Table 4.** Highest Degree Achieved by a Sample of 1,495 Graduates from Bioinformatics Program (Supplementary Data 6)

| Highest Degree | Percentage |
| --- | --- |
| Bachelor's Degree | 57.4% |
| Master's Degree | 12.9% |
| High school degree or equivalent (e.g., GED) | 7.3% |
| Some college but no degree | 11.5% |
| Less than high school degree | 2.0% |
| I prefer not to answer | 1.3% |
| Doctor of Philosophy (PhD) | 1.5% |
| Doctor of Medicine (MD) | 1.9% |
| Associate degree | 4.1% |

## References

1. Pravettoni, G., and Triberti, S. “P5 eHealth: An Agenda for the Health Technologies of the Future.”, 2020, pp. 75–90. OAPEN, library.oapen.org/bitstream/handle/20.500.12657/22850/1/1007311.pdf#page=76.

2. Gusila, I., Topa, A., Zarbailov, N., Lungu, N., Curocichin, G. (2024). Personalised Medicine Implementation in Low- and Middle-Income Countries. In: Sontea, V., Tiginyanu, I., Railean, S. (eds) 6th International Conference on Nanotechnologies and Biomedical Engineering. ICNBME 2023. IFMBE Proceedings, vol 92. Springer, Cham. 10.1007/978-3-031-42782-4_44

3. Pramesh, C.S., Badwe, R.A., Bhoo-Pathy, N. et al. Priorities for cancer research in low- and middle-income countries: a global perspective. Nat Med 28, 649–657 (2022). 10.1038/s41591-022-01738-x

4. Budd A, Corpas M, Brazas MD, Fuller JC, Goecks J, Mulder NJ, et al. (2015) A Quick Guide for Building a Successful Bioinformatics Community. PLoS Comput Biol 11(2): e1003972. 10.1371/journal.pcbi.1003972

5. Snyder, Michael P., et al. “Perspectives on ENCODE.” Nature 583.7818 (2020): 693–698.

6. Turnbaugh PJ, Ley RE, Hamady M, Fraser-Liggett CM, Knight R, Gordon JI. The human microbiome project. Nature. 2007 Oct 18;449(7164):804-10. doi: 10.1038/nature06244. PMID: 17943116; PMCID: PMC3709439. Bernstein BE, Birney E, Dunham I, Green ED, Gunter C, et al. (2012) An integrated encyclopedia of DNA elements in the human genome. Nature 489: 57–74. pmid:22955616

7. International Cancer Genome Consortium. “International network of cancer genome projects.” Nature 464.7291 (2010): 993.

8. Ramsay, M. et al. H3Africa AWI-Gen Collaborative Centre: a resource to study the interplay between genomic and environmental risk factors for cardiometabolic diseases in four sub-Saharan African countries. Glob. Health Epidemiol. Genom. 1, e20 (2016).

9. Skantharajah, Neerjah, et al. “Equity, diversity, and inclusion at the Global Alliance for Genomics and Health.” Cell genomics 3.10 (2023).

10. Choudhury, A., Aron, S., Botigué, L.R. et al. High-depth African genomes inform human migration and health. Nature 586, 741–748 (2020). 10.1038/s41586-020-2859-7

11. Sirugo G, Williams SM, Tishkoff SA. The Missing Diversity in Human Genetic Studies. Cell. 2019 Mar 21;177(1):26–31. doi: 10.1016/j.cell.2019.02.048. Erratum in: Cell. 2019 May 2;177(4):1080. doi: 10.1016/j.cell.2019.04.032. PMID: 30901543; PMCID: PMC7380073.

12. Guadagnolo B. Ashleigh, et al. “Medical mistrust and less satisfaction with health care among Native Americans presenting for cancer treatment.” Journal of health care for the poor and underserved 20.1 (2009): 210–226.

13. Pacheco, Christina M., et al. “Moving forward: breaking the cycle of mistrust between American Indians and researchers.” American journal of public health 103.12 (2013): 2152–2159.

14. Chadwick, Jennifer Q., et al. “Genomic research and American Indian tribal communities in Oklahoma: learning from past research misconduct and building future trusting partnerships.” American journal of epidemiology 188.7 (2019): 1206–1212.

15. Sanford, James A., et al. “Molecular transducers of physical activity consortium (MoTrPAC): mapping the dynamic responses to exercise.” Cell 181.7 (2020): 1464–1474.

16. Jain, Sanjay, et al. “Advances and prospects for the Human BioMolecular Atlas Program (HuBMAP).” Nature cell biology 25.8 (2023): 1089–1100.

17. Rozenblatt-Rosen, Orit, et al. “The human tumor atlas network: charting tumor transitions across space and time at single-cell resolution.” Cell 181.2 (2020): 236–249.

18. Bridge2AI https://commonfund.nih.gov/bridge2ai; Accessed 8 June,2024.

19. Peterson, Jane, et al. “The NIH human microbiome project.” Genome research 19.12 (2009): 2317–2323.

20. Lonsdale, John, et al. “The genotype-tissue expression (GTEx) project.” Nature genetics 45.6 (2013): 580–585.

21. Mollick, Ethan, and Lilach Mollick. “Assigning AI: Seven approaches for students, with prompts.” arXiv preprint arXiv:2306.10052 (2023).

22. Nazi, Kim M., et al. “Consumer health informatics: engaging and empowering patients and families.” Clinical informatics study guide: text and review (2016): 459–500.

23. Zhang, Peng, and Maged N. Kamel Boulos. “Generative AI in medicine and healthcare: Promises, opportunities and challenges.” Future Internet 15.9 (2023): 286.

24. Alowais, Shuroug A., et al. “Revolutionizing healthcare: the role of artificial intelligence in clinical practice.” BMC medical education 23.1 (2023): 689.

25. DeLozier, S. J., & Rhodes, M. G. (2017). Assessing the effectiveness of personalized learning in a large, introductory biology course. Journal of Microbiology & Biology Education, 18(1), 1–7.

26. Brown, Peter C., Henry L. Roediger III, and Mark A. McDaniel. Make it stick: The science of successful learning. Harvard University Press, 2014.

27. Dosch and Zidon. (2014). “The Course Fit Us”: Differentiated Instruction in the College Classroom. International Journal of Teaching and Learning in Higher Education. 26(3): 343–357.

28. Tomlinson, et. al. (2003). Differentiating Instruction in Response to Student Readiness, Interest, and Learning Profile in Academically Diverse Classrooms: A Review of Literature. Journal for the Education of the Gifted. 27(2–3): 119–145

29. Turner, et. al. (2017). Differentiating Instruction for Large Classes in Higher Education. International Journal of Teaching and Learning in Higher Education. 29(3): 490–500.

30. Amazon Web Services. “Architecting of HIPAA Security and Compliance on Amazon Web Services.” Accessed 8 June, 2024. https://docs.aws.amazon.com/pdfs/whitepapers/latest/architecting-hipaa-security-and-complianc e-on-aws/architecting-hipaa-security-and-compliance-on-aws.pdf#document-revisions

31. Google Cloud Platform. “HIPAA Compliance on Google Cloud”. Accessed 8 June, 2024. https://cloud.google.com/security/compliance/hipaa

32. AWS Bedrock, AWS. https://aws.amazon.com/bedrock/. Accessed 8 June, 2024.

33. Microsoft Azure OpenAI Service. https://azure.microsoft.com/en-us/products/ai-services/openai-service. Accessed 8 June, 2024.

34. GCP Vertex AI. https://cloud.google.com/vertex-ai. Accessed 8 June, 2024.

35. Yao, Yifan, et al. “A survey on large language model (llm) security and privacy: The good, the bad, and the ugly.” High-Confidence Computing (2024): 100211.

36. Li, Haoran, et al. “Privacy in Large Language Models: Attacks, Defenses and Future Directions.” arXiv, arXiv:2310.10383, 2023.

37. Zhang, Zhexin, et al. “SafetyBench: Evaluating the Safety of Large Language Models with Multiple Choice Questions.” arXiv, arXiv:2309.07045, 2023.

38. Wilkinson, Mark D., et al. “The FAIR Guiding Principles for scientific data management and stewardship.” Scientific data 3.1 (2016): 1–9.

39. Alavi, Arash, et al. “Real-time alerting system for COVID-19 and other stress events using wearable data.” Nature medicine 28.1 (2022): 175–184.

40. Dibia, V. (2023). Lida: A tool for automatic generation of grammar-agnostic visualizations and infographics using large language models. arXiv preprint arXiv:2303.02927.

41. Amazon Q in QuickSight. https://aws.amazon.com/quicksight/q/. Accessed 8 June, 2024.

42. Anthropic’s Claude 2.1 on Amazon Bedrock, Accessed 8 June, 2024. https://aws.amazon.com/about-aws/whats-new/2023/11/claude-2-1-foundation-model-anthropic-amazon-bedrock/. Accessed 8 June, 2024.

43. Anthropic’s Claude 3 Haiku on Amazon Bedrock. https://aws.amazon.com/about-aws/whats-new/2024/03/anthropics-claude-3-haiku-model-amazon-bedrock/. Accessed 8 June, 2024.

44. Anthropic’s Claude 3 Opus on Amazon Bedrock, Accessed 8 June, 2024. https://aws.amazon.com/about-aws/whats-new/2024/04/anthropics-claude-3-opus-amazon-bedrock/. Accessed 8 June, 2024.

45. Anthropic’s Claude 3 Sonnet on Amazon Bedrock, Accessed 8 June, 2024. https://aws.amazon.com/about-aws/whats-new/2024/03/anthropics-claude-3-sonnet-model-amazonbedrock/. Accessed 8 June, 2024.

46. Google, Gemini Team. Gemini 1.5-pro: Unlocking multimodal understanding across millions of tokens of context. arXiv. 10.48550/arXiv.2403.05530 (2024).

47. GPT-3.5 Turbo. OpenAI API. https://platform.openai.com/docs/models/gpt-3-5-turbo. Accessed 8 June 2024.

48. GPT-4 Turbo and GPT-4. OpenAI API. https://platform.openai.com/docs/models/gpt-4-turbo-and-gpt-4. Accessed 8 June 2024.

49. GPT-4o. OpenAI API. https://platform.openai.com/docs/models/gpt-4o. Accessed 8 June 2024.

50. Meta’s Llama 2 model 70B on Amazon Bedrock. https://aws.amazon.com/about-aws/whats-new/2023/11/llama-2-70b-foundation-model-meta-amazon-bedrock/. Accessed 8 June,2024.

51. Chen, L., Zaharia, M., and J. Zou. “How Is ChatGPT’s Behavior Changing over Time?” Harvard Data Science Review, vol. 6, 2024

52. Mishra, Tejaswini, et al. “Pre-symptomatic detection of COVID-19 from smartwatch data.” Nature biomedical engineering 4.12 (2020): 1208–1220.

53. Zhou, Wenyu, et al. “Longitudinal multi-omics of host–microbe dynamics in prediabetes.” Nature 569.7758 (2019): 663–671.

54. Schüssler-Fiorenza Rose, Sophia Miryam, et al. “A longitudinal big data approach for precision health.” Nature medicine 25.5 (2019): 792–804.

55. Contrepois, Kévin, et al. “Molecular choreography of acute exercise.” Cell 181.5 (2020): 1112–1130.

56. Cummings, M. P., & Temple, G. G. (2010). Broader incorporation of bioinformatics in education: opportunities and challenges. Briefings in bioinformatics, 11(6), 537–543.

57. Ranganathan, S. (2005). Bioinformatics education—perspectives and challenges. PLoS computational biology, 1(6), e52.

58. Bertero, L., & Tuvé, C. (2021). Biomedical Big Data: Challenges and Opportunities. In M. Dashti (Ed.), Data Science, Artificial Intelligence and Machine Learning Applications (pp. 135–158). Springer.

59. Nagarajan, M., Verma, A., & McCarroll, R. (2020). Use of Cloud Computing Technologies for Biomedical Data Analysis. In Handbook of Big Data Technologies (pp. 1-25). Springer.

60. Kluyver, Thomas, et al. “Jupyter Notebooks-a publishing format for reproducible computational workflows.” Elpub 2016 (2016): 87–90.

61. Bahmani, A., Sedigh, S., & Hurson, A. (2012). Ontology-based recommendation algorithms for personalized education. In Database and Expert Systems Applications: 23rd International Conference, DEXA 2012, Vienna, Austria, September 3-6, 2012. Proceedings, Part II 23 (pp. 111–120). Springer Berlin Heidelberg.

62. Mengstie, B. Impact of microfinance on women’s economic empowerment. J Innov Entrep 11, 55 (2022). 10.1186/s13731-022-00250-3

63. UNICEF. “Girls’ Education.” UNICEF, https://www.unicef.org/education/girls-education.

64. Duncan, A., Premnazeer, M. & Sithamparanathan, G. Massive open online course adoption amongst newly graduated health care providers. Adv in Health Sci Educ 27, 919–930 (2022). 10.1007/s10459-022-10113-x

65. Fu, Q., Gao, Z., Zhou, J., & Zheng, Y. (2021). CLSA: A novel deep learning model for MOOC dropout prediction. Computers & Electrical Engineering, 94, 107315. 10.1016/j.compeleceng.2021.107315

66. Gütl, Christian, et al. “Attrition in MOOC: Lessons learned from drop-out students.” Learning Technology for Education in Cloud. MOOC and Big Data: Third International Workshop, LTEC 2014, Santiago, Chile, September 2-5, 2014. Proceedings 3. Springer International Publishing, 2014.

67. Alhadabi A. Science Interest, Utility, Self-Efficacy, Identity, and Science Achievement Among High School Students: An Application of SEM Tree. Front Psychol. 2021 Sep 9;12:634120. doi: 10.3389/fpsyg.2021.634120. PMID: 34566743; PMCID: PMC8458621.

68. Honicke T., Broadbent J. (2016). The influence of academic self-efficacy on academic performance: a systematic review. Educ. Res. Rev. 17, 63–84. 10.1016/j.edurev.2015.11.002

69. Stets J., Brenner P., Burke P., Serpe R. (2017). The science identity and entering a science occupation. Soc. Sci. Res. 64, 1–14. 10.1016/j.ssresearch.2016.10.016

70. Kirbulut Z., Uzuntiryaki-Kondakci E. (2018). Examining the mediating effect of science self-efficacy on the relationship between metavariables and science achievement. Int. J. Sci. Educ. 41, 995–1014. 10.1080/09500693.2019.1585594

71. Kobicheva, A. et al. Students’ affective learning outcomes and academic performance in the blended environment at university: comparative study. Sustainability 14, 11341; 10.3390/su141811341 (2022).

72. Hornbæk, Kasper, and Hertzum, Morten. “Technology Acceptance and User Experience: A Review of the Experiential Component in HCI.” ACM Transactions on Computer-Human Interaction, vol. 24, no. 5, 2017, pp. 1–30, doi:10.1145/3127358.

73. Karan Girotra, Lennart Meincke, Christian Terwiesch, Karl T. Ulrich (July 2023): Ideas are Dimes a Dozen: Large Language Models for Idea Generation in Innovation, https://papers.ssrn.com/sol3/papers.cfm?abstract_id=4526071

74. Guzik, Erik E., Christian Byrge, and Christian Gilde. “The originality of machines: AI takes the Torrance Test.” Journal of Creativity 33.3 (2023): 100065.

75. Peng, Sida, et al. “The impact of ai on developer productivity: Evidence from github copilot.” arXiv preprint arXiv:2302.06590 (2023).

76. Cormen, Thomas H., et al. Introduction to algorithms. MIT press, 2022.

77. Doleck, T., Bazelais, P., Lemay, D. J., Saxena, A., & Basnet, R. B. (2017). Algorithmic thinking, cooperativity, creativity, critical thinking, and problem solving: Exploring the relationship between computational thinking skills and academic performance. Journal of Computers in Education, 4(4), 355–369.

78. Riedemann, L., Labonne, M. & Gilbert, S. The path forward for large language models in medicine is open. npj Digit. Med. 7, 339 (2024).

79. 1000 Genomes Project Consortium. “A global reference for human genetic variation.” Nature 526.7571 (2015): 68.

80. Forbes, S., et al. “COSMIC 2005.” British journal of cancer 94.2 (2006): 318–322.

81. Hickey, John W., et al. “Organization of the human intestine at single-cell resolution.” Nature 619.7970 (2023): 572–584.

82. McNamara, Katherine L., et al. “Spatial proteomic characterization of HER2-positive breast tumors through neoadjuvant therapy predicts response.” Nature cancer 2.4 (2021): 400–413.

83. Galeano Niño, Jorge Luis, et al. “Effect of the intratumoral microbiota on spatial and cellular heterogeneity in cancer.” Nature 611.7937 (2022): 810–817.

84. Boettiger, Carl. “An introduction to Docker for reproducible research.” ACM SIGOPS Operating Systems Review 49.1 (2015): 71–79.

85. Papin, Jason A., et al. “Improving reproducibility in computational biology research.” PLOS Computational Biology 16.5 (2020): e1007881.

86. Wu, Sunny Z., et al. “A single-cell and spatially resolved atlas of human breast cancers.” Nature genetics 53.9 (2021): 1334–1347.

87. Wang, J., Fan, W. The effect of ChatGPT on students’ learning performance, learning perception, and higher-order thinking: insights from a meta-analysis. Humanit Soc Sci Commun 12, 621 (2025).

88. Liffiton, Mark, et al. “Codehelp: Using large language models with guardrails for scalable support in programming classes.” Proceedings of the 23rd Koli Calling International Conference on Computing Education Research. 2023.

89. Riedemann, L., Labonne, M. & Gilbert, S. The path forward for large language models in medicine is open. npj Digit. Med. 7, 339 (2024).

